# Mutational landscape and dominant lineages in the SARS-CoV-2 infections in the state of Telangana, India

**DOI:** 10.1101/2020.08.24.20180810

**Authors:** Asmita Gupta, Radhakrishnan Sabarinathan, Pratyusha Bala, Vinay Donipadi, Divya Vashisht, Madhumohan Rao Katika, Manohar Kandakatla, Debashis Mitra, Ashwin Dalal, Murali Dharan Bashyam

## Abstract

The novel <u>S</u>evere <u>A</u>cute <u>R</u>espiratory <u>S</u>yndrome <u>C</u>oronavirus 2 (SARS-CoV-2) causing COVID-19 has rapidly turned into a pandemic, infecting millions and causing ~7 million deaths across the globe. In addition to studying the mode of transmission and evasion of host immune system, analysing the viral mutational landscape constitutes an area under active research. The latter is expected to impart knowledge on the emergence of different clades, subclades, viral protein functions and protein-protein and protein–RNA interactions during replication/transcription cycle of virus and response to host immune checkpoints. In this study we have attempted to bring forth the viral genomic variants defining the major clade(s) as identified from samples collected from the state of Telangana, India.

## Introduction

The outbreak of COVID-19 caused by the <u>S</u>evere <u>A</u>cute <u>R</u>espiratory <u>S</u>yndrome <u>C</u>orona<u>V</u>irus 2 (SARS-CoV-2) in the Hubei province of China during late December 2019, has since taken the shape of a global pandemic, spreading across more than 200 countries, and resulting in approximately 7,75,581 deaths worldwide, as per recent statistics by World Health Organization (WHO) (https://covid19.who.int/, August 19, 2020). SARS-CoV-2 belongs to the subfamily *Coronavirinae* of the *Coronaviridae* family, classified under the order *Nidovirales*^1^. After host entry, the 29.9 kb positive sense, single-stranded, unsegmented RNA genome of this virus gives rise to 15 non-structural proteins (nsps, 1-15). The subsequent replication and transcription cycles produce a genomic RNA template along with nine subgenomic RNAs, which are further translated to form major structural proteins viz.Spike (S), Envelope (E), Nucleocapsid (N) and Membrane (M) proteins ^2^. They share a high degree of sequence similarity with Severe Acute Respiratory Syndrome coronavirus (SARS-CoV, >80%) and a moderate sequence similarity with Middle East Respiratory Syndrome coronavirus (MERS-COV, >50%)^3^. The severity of the pandemic has demanded a concerted effort to comprehensively study the actively changing mutational landscape of the virus across multiple demographic locations and identify potential variant clusters or individual variants which might play a critical role in its spread and transmission dynamics.

The state of Telangana, located in south-central India, has seen an unusually high rate of infection and there appears to have been a sharp spike in the number of cases beginning from the second half of April 2020. In this study, we aimed at identifying the dominant lineages present in the samples collected in Telangana, among all identified lineages of SARS-CoV-2. We further endeavoured to draft a comprehensive mutational landscape of the viral genome from among patient samples in Telangana state. Towards this end, we applied next generation sequencing to determine the complete sequence of 210 SARS-CoV-2 RNA samples.

## Data and Methods

### Sample collection and processing

The Centre for DNA Fingerprinting and Diagnostics (CDFD), Hyderabad, initiated reverse transcription-PCR (RT-PCR) based diagnostics for Covid-19 infection after approvals from Secretary, Department of Biotechnology (DBT), Government of India, Indian Council of Medical Research (ICMR) nominated nodal officer in Hyderabad, Telangana as well as from the Telangana state government. The work was initiated following approvals from the Institutional Bioethics committee and Biosafety committee. The samples were obtained as nasopharyngeal swabs collected in Viral Transport Medium from different parts of Telangana from patients with symptoms suggestive of Covid-19 as well as asymptomatic primary contacts of affected cases. The samples were transported to CDFD within 24 hours while maintaining a cold chain. Total RNA was isolated using the RNA isolation kit as per manufacturer’s instructions (QIAmp Viral RNA Mini Kit; Cat #52906; Qiagen, Hilden, Germany). Each RNA sample was subjected to RT-PCR for multiple viral genes (including E-gene and RDRP gene) using the LabGun COVID-19 assay (Cat #CV9017B; LabGenomics, Republic of Korea) or the Allplex 2019-nCoV Assay (Cat #RP10250X, Seegene, Republic of Korea). The samples which tested positive in RT-PCR analysis were included for viral genome sequencing. Since RDRP consistently provided more robust amplification than E-gene and is a SARS-COVID-2 specific gene (unlike E-gene which is specific for all respiratory coronaviruses), we considered Ct values of RDRP gene alone for analysis. Samples exhibiting an RDRP Ct value greater than 10 and less than 35 were chosen for sequencing.

### Sequencing protocol

Sequencing of SARS-CoV-2 RNA samples was performed using protocol described earlier (Quick et al., 2020) with slight modifications. Briefly, RNA isolated from nasopharyngeal swabs was reverse transcribed using random primer mix (New England Biolabs, Massachusetts, United States), and Superscript-IV (Thermofisher Scientific, Massachusetts, United States). The resulting cDNA was subjected to a 3-step multiplex PCR using nCoV-2019/V3 primer pools (Eurofins, India) 1, 2 and 3. The ~400 bp amplicons thus obtained in the pools were combined, purified using Agencourt AMPure XP beads (Beckman Coulter, California, United States) and eluted in 45μl elution buffer (Qiagen, Hilden, Germany). DNA libraries for Illumina sequencing were prepared using the NEB Next Ultra II DNA Library Prep Kit for Illumina (New England Biolabs, Massachusetts, United States), according to the manufacturer’s protocol. Paired-End Sequencing (2×250 bp) was performed on the Miseq FGx (Illumina Inc, California, United States) with a targeted depth of 0.5 million reads per sample (~4,000X coverage). Libraries for Nanopore sequencing were prepared using the Ligation sequencing kit (LSK-109; Oxford Nanopore Technologies, London, United Kingdom). Barcoded libraries were pooled (12-24 samples each) and sequenced on a MinION flow cell in GridION (Oxford Nanopore Technologies, London, United Kingdom). Sequencing was performed with a targeted depth of 0.1 million reads per sample (up to 24 hours).

### Mutation analysis

All raw fastq files from Illumina were checked for overall sequencing quality, presence of adapters and bad quality reads using FastQC and Fastp^4^. The adapter sequences were trimmed using a wrapper script for Cutadapt^5^ tool, called Trim Galore. The filtered reads were aligned to the reference strain NC_045512.1, Severe acute respiratory syndrome coronavirus 2 isolate Wuhan-Hu-1, using bwa-mem^6^ algorithm with default parameters. Mapping quality was assessed using samtools^7^ and BAMStats. Post alignment, the reads were filtered, sorted and indexed using samtools, and any primer sequences were masked using iVar^8^. Subsequent mutation calling and generation of consensus sequence was performed using samtools mpileup and iVar (https://github.com/connor-lab/ncov2019-artic-nf/). The resulting VCF files were annotated using snpEff^9^. For processing the nanopore data, we followed the protocol suggested by ARTIC pipeline (https://github.com/artic-network/fieldbioinformatics, https://github.com/connor-lab/ncov2019-artic-nf/) for mutation calling as well as for assembling the reads for generating the consensus sequence. A schematic describing the entire workflow is shown in **Supplementary Fig S1**. Before analysing the obtained calls, we filtered all the problematic sites prone to errors by multiple sources as recommended by De Maio et al (https://virological.org/t/issues-with-sars-cov-2-sequencing-data/473).

### Phylogenetic analysis

The consensus fasta files generated for both Illumina and Nanopore data were subjected for phylogenetic analysis using the Nextstrain pipeline with recommended default criteria for filtering, multiple sequence alignment (MSA) and nucleotide substitution calculations. To briefly summarize the workflow of the pipeline, all the consensus sequences having length < 27000 and Ns > 5% were filtered out. Three samples were removed from further analysis as their sequence data included Ns > 5%; thus all analyses were conducted on 207 individual patient viral genome sequences. A compendium of problematic sites as used earlier (https://virological.org/t/issues-with-sars-cov-2-sequencing-data/473), was also provided to mask those sites prior to MSA by MAFFT^10^. Following MSA, the workflow constructed a time-resolved phylogenetic tree using the maximum likelihood based method IQ-TREE^11^. The resultant tree was pruned and internal nodes and ancestral traits were inferred from the dates of the sample collection using TreeTime^12^. The final tree in Newick format was then customized for visualization using iTol^13^.

## Results

### General sample features

Our dataset consists of samples collected during late March to July, 2020. Interestingly, samples collected from late May till July represented a higher proportion of asymptomatic cases when compared to samples collected earlier (**Figure 1a**). A majority of our samples belonged to age group between15-62 years, with males (61%) dominating the profile distribution over females (39%) (**Figure 1b**). We also compared the distribution of cases with respect to Ct values, the latter being a proxy for viral load. Symptomatic cases appeared to be associated with higher Ct values (thus lower viral load), compared to asymptomatic ones, which was unexpected (**Figure 1c**). there was a reduction in the Ct values in samples, as we neared the end of June, 2020, implying that more recent samples seemed to carry a higher viral load than earlier samples (**Supplementary figure S2a**). The distribution of Ct values with respect to age showed no clear correlation (**Supplementary figure S2b**). However, there was a consistent rise in the fraction of samples showing asymptomatic behaviour, as calculated from a cumulative increment, as opposed to symptomatic, starting from the end of May (**Figure 1d**). The locale distribution of the samples in and around the district of Hyderabad is indicated in **Supplementary figure S2c**.

**Figure 1:**
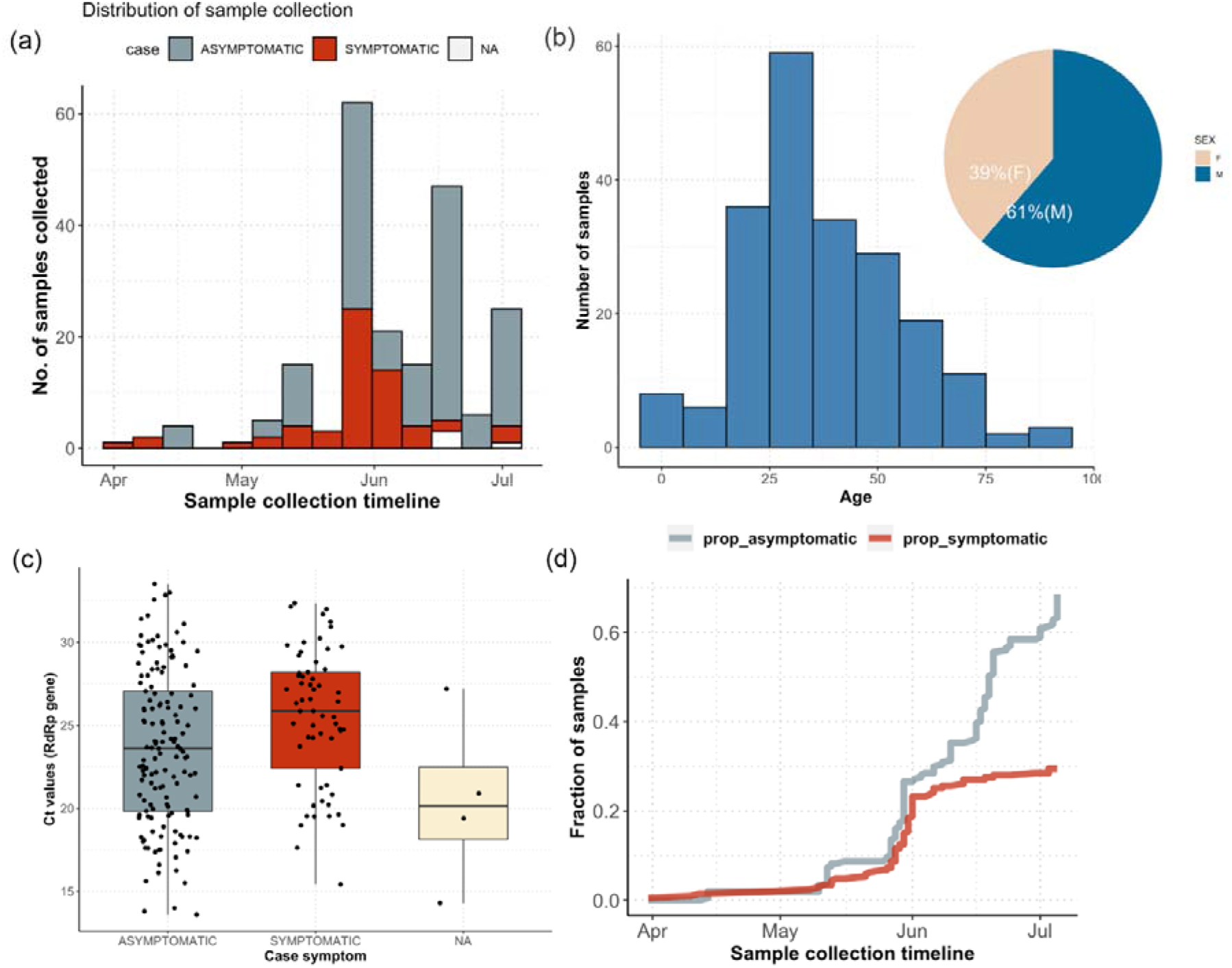
Characteristics of the dataset used in this study. (a) Distribution of sample collection timeline (b) age and gender distribution across samples (c) distribution of symptoms with respect to Ct values of RdRp gene (nsp12). (d) cumulative increment of symptomatic and asymptomatic fraction of samples along the sample collection timeline.

### Phylodynamic epidemiological clustering of SARS-Cov2 strains

The phylodynamic clustering of samples suggested presence of 2 minor (19A, 20A) and one major clade (20B). The phylogenetic clustering was rooted with respect to the sequence from the Wuhan Wu-1 strain (NC_045512.1), which has been annotated as the base clade 19A. One sample (collected in April, 2020) seemed to directly emerge from this base clade. Further, five samples harbored the C13730T mutation linking them to the base clade (establishing sub-clade 19A/C13730T). This split was marked by missense mutations G1820A (ORF1a, G519S), C6310A (ORF1a, S2015R), C6312A (ORF1a, T2016K), C28311T (N, P13L), and two synonymous mutations C19524T (ORF1a, L6425L) and C23929T (S, Y789Y). The first major split in our dataset was observed at around the starting of April, marking the onset of 20A clade, and was characterized by the appearance of co-occurring mutations C241T (5’-UTR), synonymous C3037T (ORF1ab, F924F), and missense C14408T (ORF1ab, P4720L) and A23403G (S, D614G). Seven samples belonged to this 20A clade, directly linked to the dominant viral lineage established in Europe (Belgium, Italy, of which one exhibited a distinct 20A/18877T profile. This division retained none of the mutations found in the initial samples belonging to 19A and 19A/C13730T cluster (**Supplementary figure S3**). From the middle of April and onwards, the profile was strongly dominated by the 20B clade, which also formed the second cluster within this division, with a characteristic mutation of GGG28881/28882/28883>AAC (**Figure 2 and Supplementary figure S4**. Thus, of all the clades identified, our dataset was strongly populated by the presence of a single major clade 20B. From a time resolved mutational map calculated for all samples (**Supplementary figure S4**), we observed that the more recent samples, collected from the end of June onwards, did not show many of the mutations found in ORF1a, C5700A (nsp3, A1812D), C6573T (nsp3, S2103F) and C25528T (ORF3a, L46F). These samples also coincided with the most diverged samples along the phylogenetic time-tree, indicating towards a newer divergence path of the virus among the later infections. We checked whether these samples belonged to any single cluster within the phylogenetic tree, but they were found to be interspersed among different branches (**Figure 2, Supplementary figure S4**). Within the dominant 20B clade, a major proportion of samples between the age group of 15 and 50 were found to be asymptomatic, while the symptomatic patients mainly belonged to samples collected from age group >50. As also discussed before, the viral load, calculated in terms of RdRp gene, was found to be primarily associated with asymptomatic patients. This behaviour can explain the higher transmission rates of the virus among the Indian population, in general, as opposed to per capita mortality rates, which will be further elaborated upon in the next section. A location wise distribution indicating the neighbourhood origin of the samples in the phylogeny tree has been shown in **Supplementary figure S5**.

**Figure 2:**
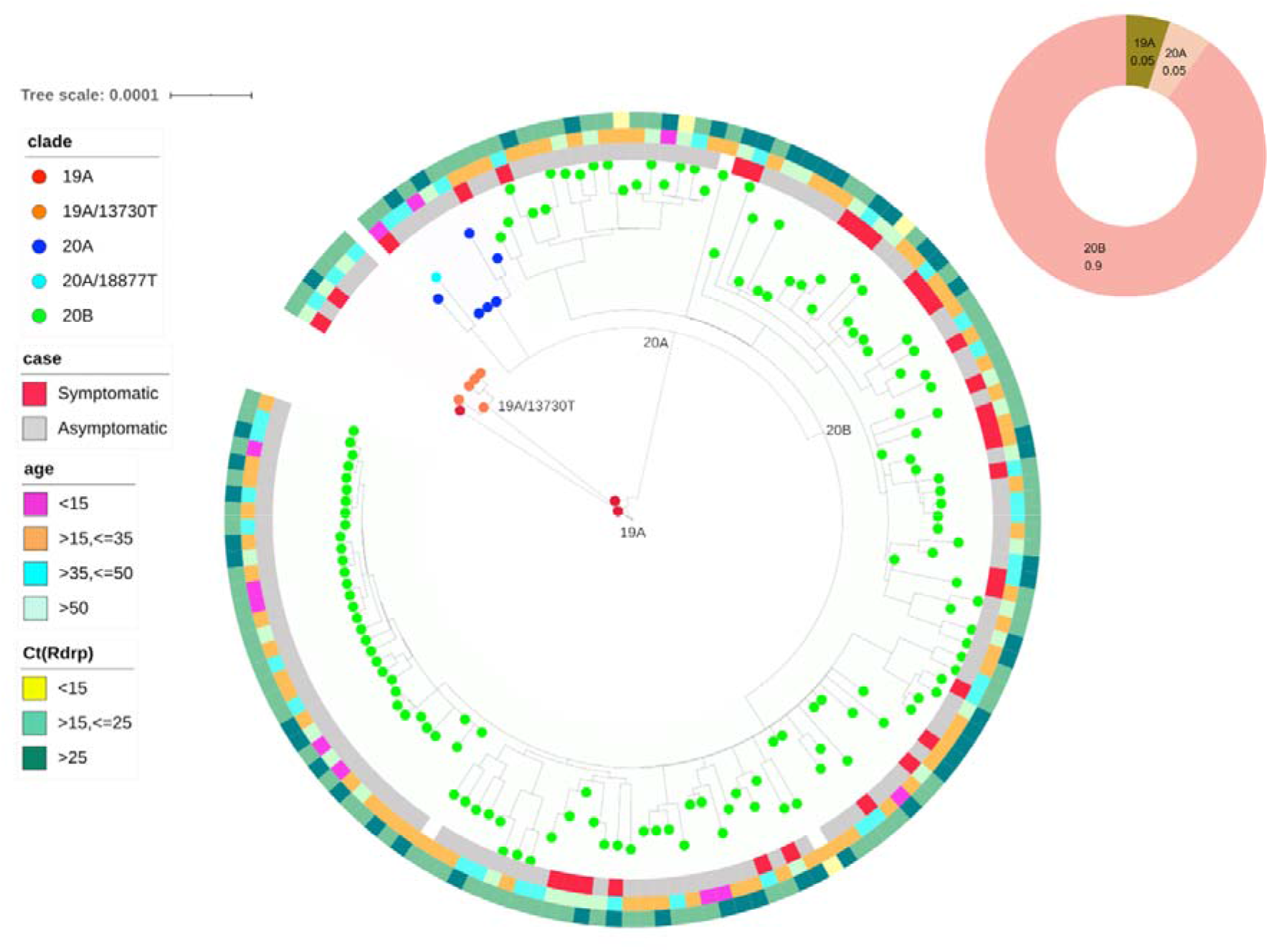
Phylodynamic tree of the samples as analyzed using Nextstrain (a) Time resolved phylogenetic tree representing all 205 samples which passed filtering criteria during Nextstrain analysis. (b) Distribution of clades across the dataset.

We performed Nextstrain analysis separately on our Illumina and nanopore datasets, in order to rule out the possibility of incorporation of any bias in cluster formation due to the sequencing platform. The clusters obtained did not show any segregation due to the platform used and were distributed across the clades (data not shown).

### Mutational landscape of SARS-CoV-2

From the mutation analysis on the filtered, combined pool of 207 sequences from Illumina and Nanopore data, we obtained a total 302 mutations across the SARS-CoV-2 genome (**Supplementary data D1**). From this set, 17 mutations were consistently found to be present in >10% of samples (**Figure 3a**). The proportion of asymptomatic cases for each of these 17 high frequency mutations, as a fraction of the total number of cases, was found to be higher than symptomatic cases (**Figure 3b**). Further, for each of the 17 high frequency mutation identified in this set, we calculated the cumulative increment in the frequency of case symptoms as a function of time and noted a consistent increase in the frequency of asymptomatic cases specially for missense mutations (**Supplementary figure S6**). Of these 17 high frequency mutations, 5 viz. C241T (5’-UTR), C3037T (ORF1a), C14408T (nsp12, ORF1ab), A23403G (S), GGG28881-3AAC (N) were highly recurrent (present in >80% of the samples; **Figure 3a**). The A23403G (D614G) mutation in Spike protein was identified in samples as early as beginning of April, 2020. Although a highly recurrent mutation in multiple demographics, no clear correlation has been established between D614G mutation and severity of disease ^14^. The D614G mutation has almost invariably been found to be associated with C241>T, C3037>T (a silent mutation) and a mutation in RNA dependent RNA polymerase gene, nsp12 C14408>T as has also been reported earlier ^15^. The haplotype defined by the co-occurrence of these 4 mutations is the current dominant form circulating across the world. Apart from these four mutations, the nsp3 protein region within ORF1a also displayed a higher frequency of mutations, G4354A, A4372G, C5700A, C6027T, C6573T, the latter three being missense mutations. Of these, C5700 along with a silent C313T mutation has been reported to co-occur in samples collected from Western state of Maharashtra, India ^16^. The nsp4 and nsp5 proteins each harbored one high frequency missense mutation each namely C9693T and C10815T, respectively. Similarly, the ORF3a region possessed one high frequency missense mutation C25528T.

**Figure 3:**
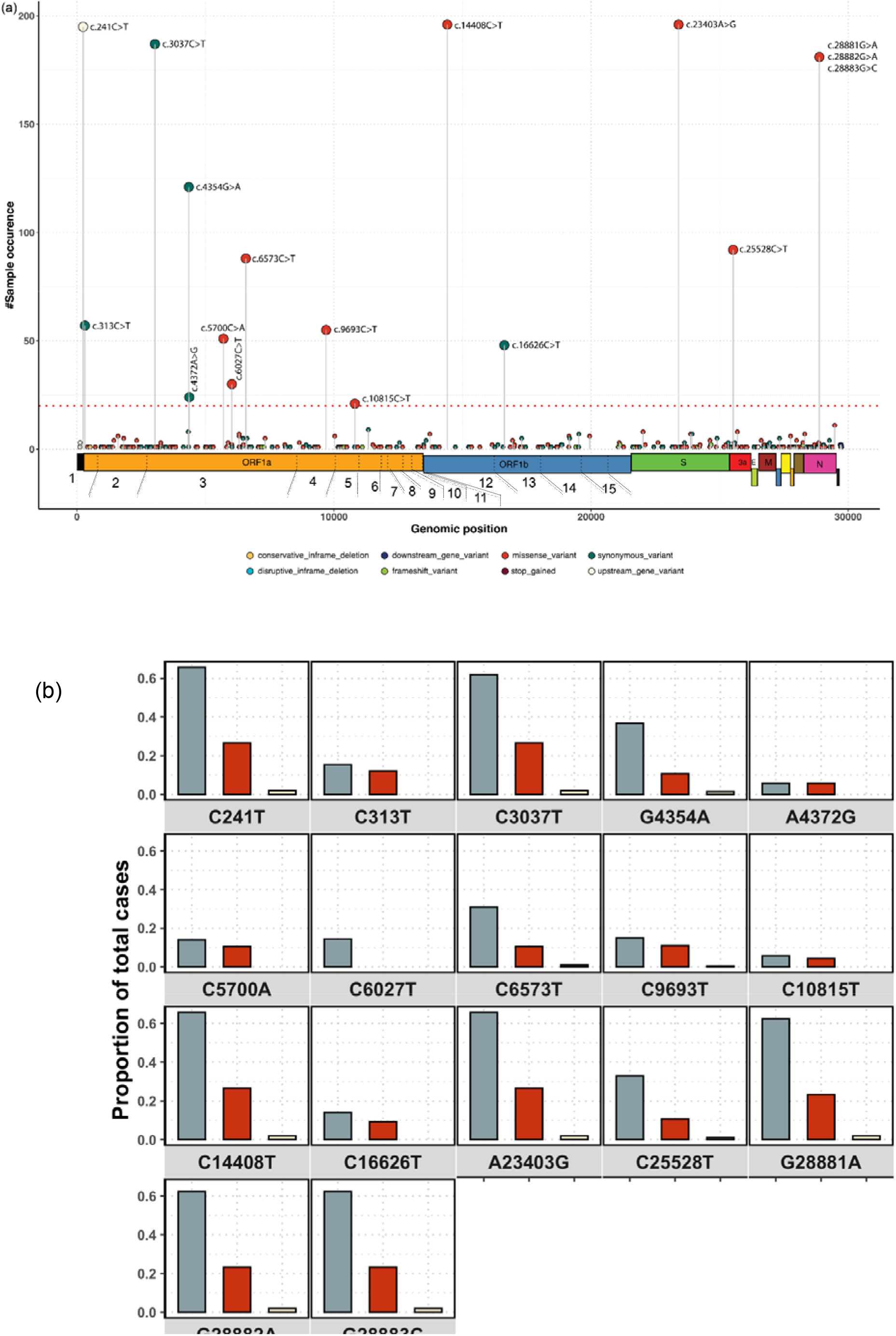
Distribution of all high frequency mutations as called on Illumina and nanopore sequencing data (a) genomic locations of all high frequency mutations (samples > 10%, indicated by red, dashed horizontal line) (b) case distribution of sample with respect to all indicated mutations. An associated cumulative frequency of cases with respect to mutations is provided in **Supplementary figure S6**.

## Discussion

In the current study, we have presented a comprehensive map of the mutations identified from the confirmed COVID-19 cases collected from the southern state of India, Telangana. After a slow progress of the outbreak during the months of February-April, the state has been witnessing a constant upsurge in the number of infections and has been listed as one of the worst affected states in the country. Identifying the mutations from samples collected over a period of time, provides a way to assess the genomic diversity which the virus might have experienced during infection and transmission. With these aspects in our purview, we have attempted to characterize the genomic epidemiology of novel coronavirus and a comprehensive mutational landscape, using a dataset of 210 samples sequenced using both Illumina and Nanopore sequencing technologies. Most of our samples were collected from mid-March with the months of June-July reporting highest collections. A significantly higher association of samples with asymptomatic behaviour was noted, which were also associated with lower Ct values. We also observed an upsurge in the asymptomatic cases compared to symptomatic. A majority of our samples belonged to the 20B clade, with the clade seeming to appear in the beginning of April. Although we did not see a clear correlation between age and viral load, it was observed that samples collected from higher age groups frequently displayed symptomatic behaviour.

More importantly, mutational analysis revealed the presence of unique mutations in the samples from Telangana, especially in the nsp3, nsp4, nsp5 and ORF3a. The nsp3 is a papain-like protease (PLP2) and nsp5 is a 3C-like protease (3CLpro), both required for cleavage of polyproteins pp1a and pp1ab to generate 16 non-structural proteins (nsp1-16) ^17^. Nsp3 is the largest multidomain protein encoded by SARS-CoV-2, and binds to viral RNA and nucleocapsid protein ^18^. The mutations identified in nsp3 viz. C6027T (P1921L), C5700A (Ala1812Asp) and C6573T (S2103F) lie in the protease like (PLP2) and protease like non-canonical (PLnc) domains of the protein. The PLnc domain has been shown to interact with a large number of nsp partners during viral replication and transcription cycle and acts as a scaffold for membrane associated replication/transcription complex (RTC) formation. The formation of RTC also requires nsp3 association with nsp4 and nsp5 ^19^. Hence, it could be speculated that these mutations can potentially alter the balance or stoichiometry of the complex formation, perhaps causing viral RTC reprogramming. The RTC complexes in coronaviruses have been shown to perform multiple tasks during replication and transcription by being able to form compositionally diverse complexes, each specific to carry out a particular process. It is possible that these mutations could bring about a switch in this compositional programming ^19^.

SARS-CoV-2 ORF3a encodes an accessory, transmembrane, ion-channel protein, belonging to the class of proteins called Viroporins, which induce innate immune signalling receptor NLRP3, leading to production of cytokines IL-1β, IL-6 and tumor necrosis factor (TNF) ^20^. This often results in tissue inflammation in a SARS-CoV-2 infection. The most common mutations identified in ORF3a among the prevalent lineages across the world is Q57H and G251V ^21^. However, the C25528T (L46F) mutation observed in this study localizes to the same domain I as Q57H, in the N-terminal signal-peptide region of the protein. This region is critical for viral localization to the Golgi apparatus. Functional impact of these mutations on the location, and subsequent translation and assembly, might require additional experimental evidences.

In addition to these high frequency mutations, we also observed a few low frequency mutations in the Spike, N, nsp3 and nsp2 proteins. A detailed study of these mutations and their functional impact on the viral life cycle is currently underway.

In conclusion, we report the comprehensive mutation landscape of more than 200 individual SARS-CoV-2 viral genome sequences isolated from COVID-19 patients or primary contacts. We report the 20B clade as the major clade in Telangana which is similar to reports from West India. The data forms a starting point for the state government machinery to conduct further studies on virus transmission helping in taking informed public health decisions. The genomic mutations also inform on potential mechanisms being employed during evasion of the host’s immune response and traces of higher or lower pathogenicity, if any, being developed over the span of time, thus significantly impacting efforts in vaccine development.

## Data Availability

The sequence data is being deposited into the GISAID database

## Acknowledgements

We are thankful to Drs Rashna Bhandari and R Harinarayanan, CDFD, Hyderabad, for co-ordinating the establishment of the COVID-19 testing laboratory at CDFD. All volunteers and ‘COVID warriors’ from CDFD, Hyderabad, are gratefully acknowledge for their significant contribution in screening of samples. We are grateful to all Government district hospitals in the Telangana State for facilitating screening of suspected COVID-19 patients. We also thank the Telangana State Government, the Indian Council of Medical Research, Government of India, the University of Hyderabad, Hyderabad and CDFD, Hyderabad, for procurement of consumables and equipment to perform screening of patient samples and the Department of Biotechnology, Government of India for financial support through the National Genomics Core project (BT/INF/22/SP28169/2019,07/03/2019). R.S. acknowledges funding support from NCBS-TIFR and Ramanujan Fellowship (SB/S2/RJN-071/2018). A.G. acknowledges funding support from Science and Engineering Research Board, Department of Science and Technology (DST-SERB) in the form of National-Postdoctoral Fellowship (N-PDF).

## Supplementary data

**Supplementary figure S1:**
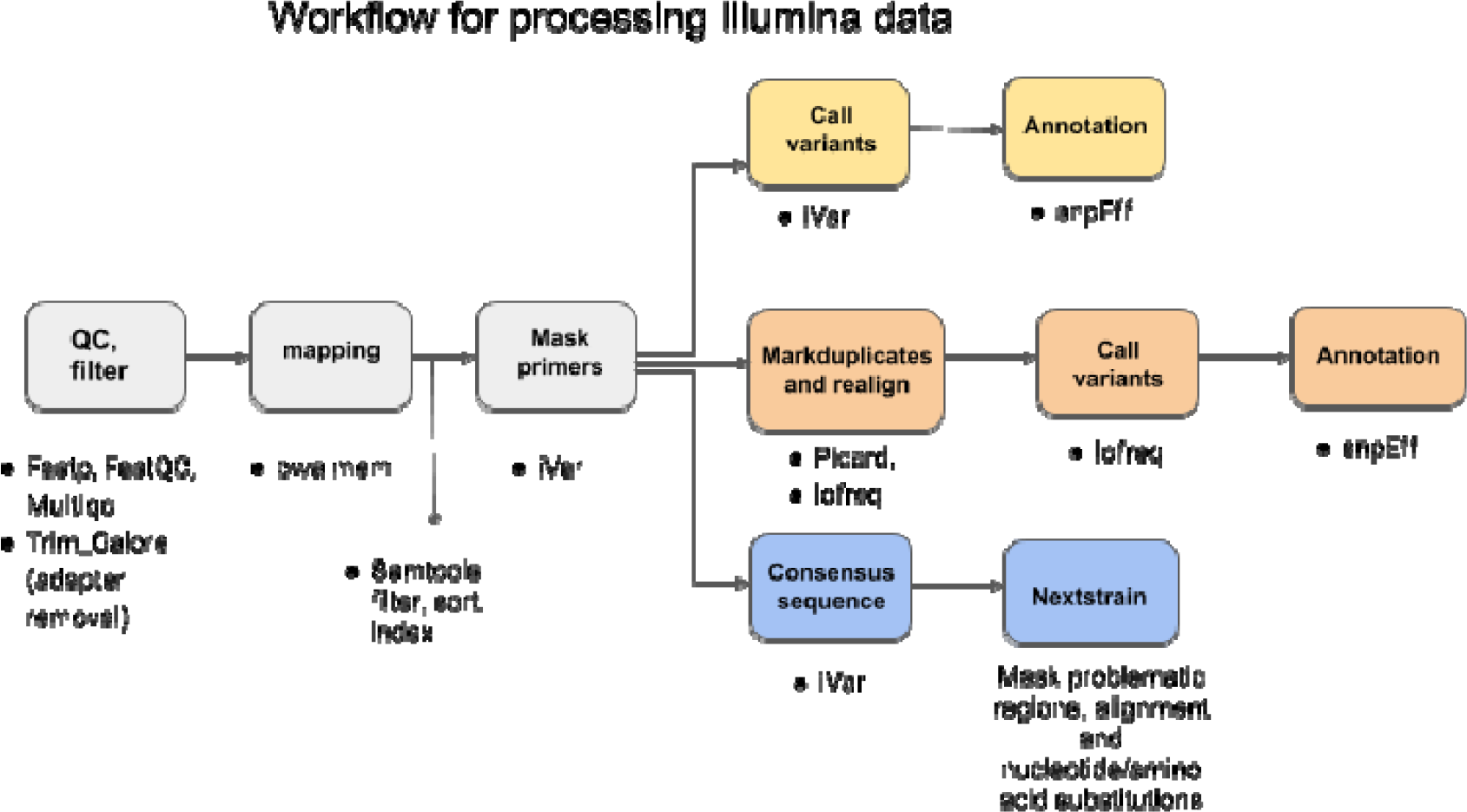
Detailed workflow of the methodology and various stages.

**Supplementary figure S2:**
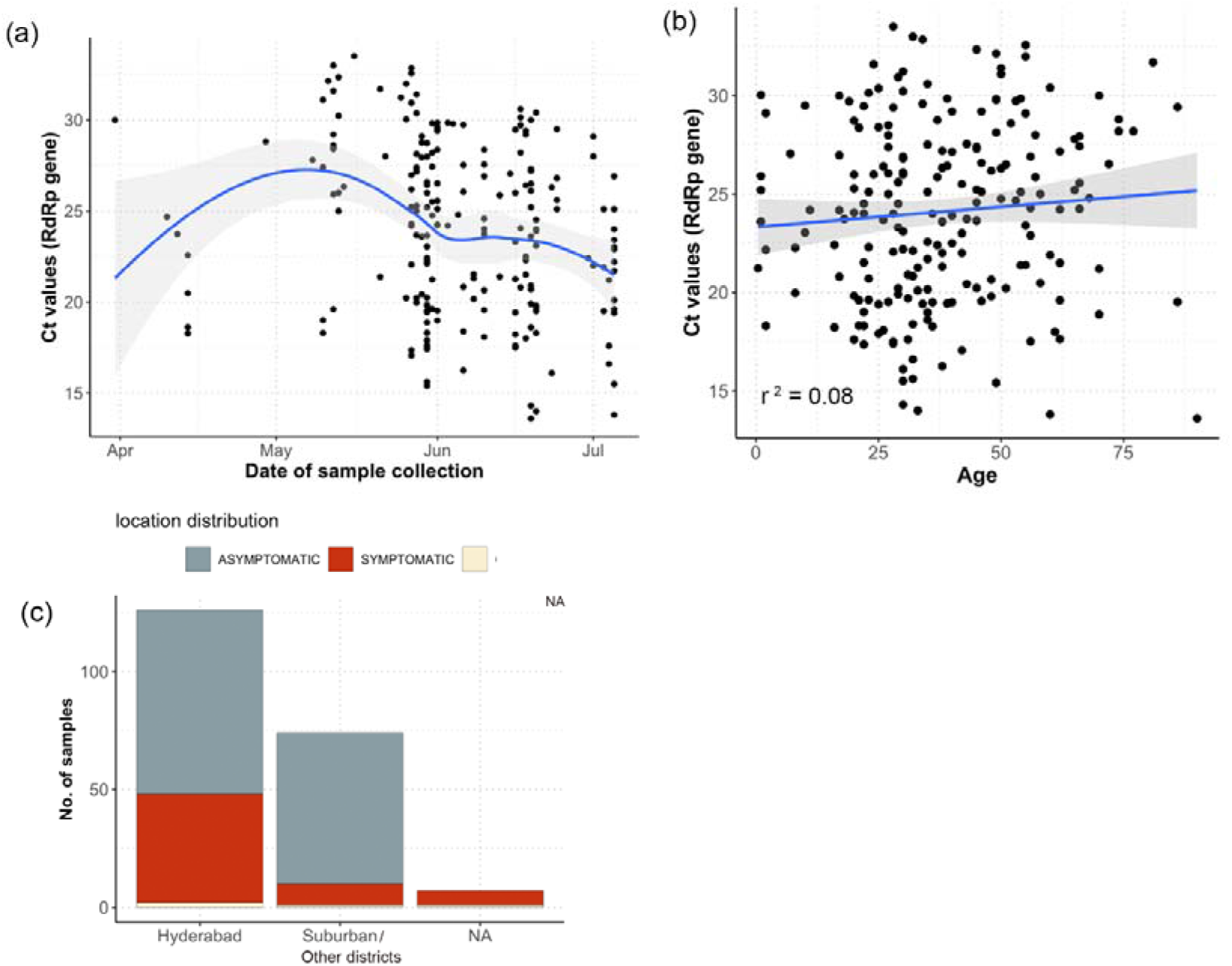
Sample features: (a) Ct value distribution of samples with respect to date of sample collection (b) Correlation between age and Ct values of the samples (c) Distribution of samples with respect to locations within the district of Hyderabad.

**Supplementary figure S3:**
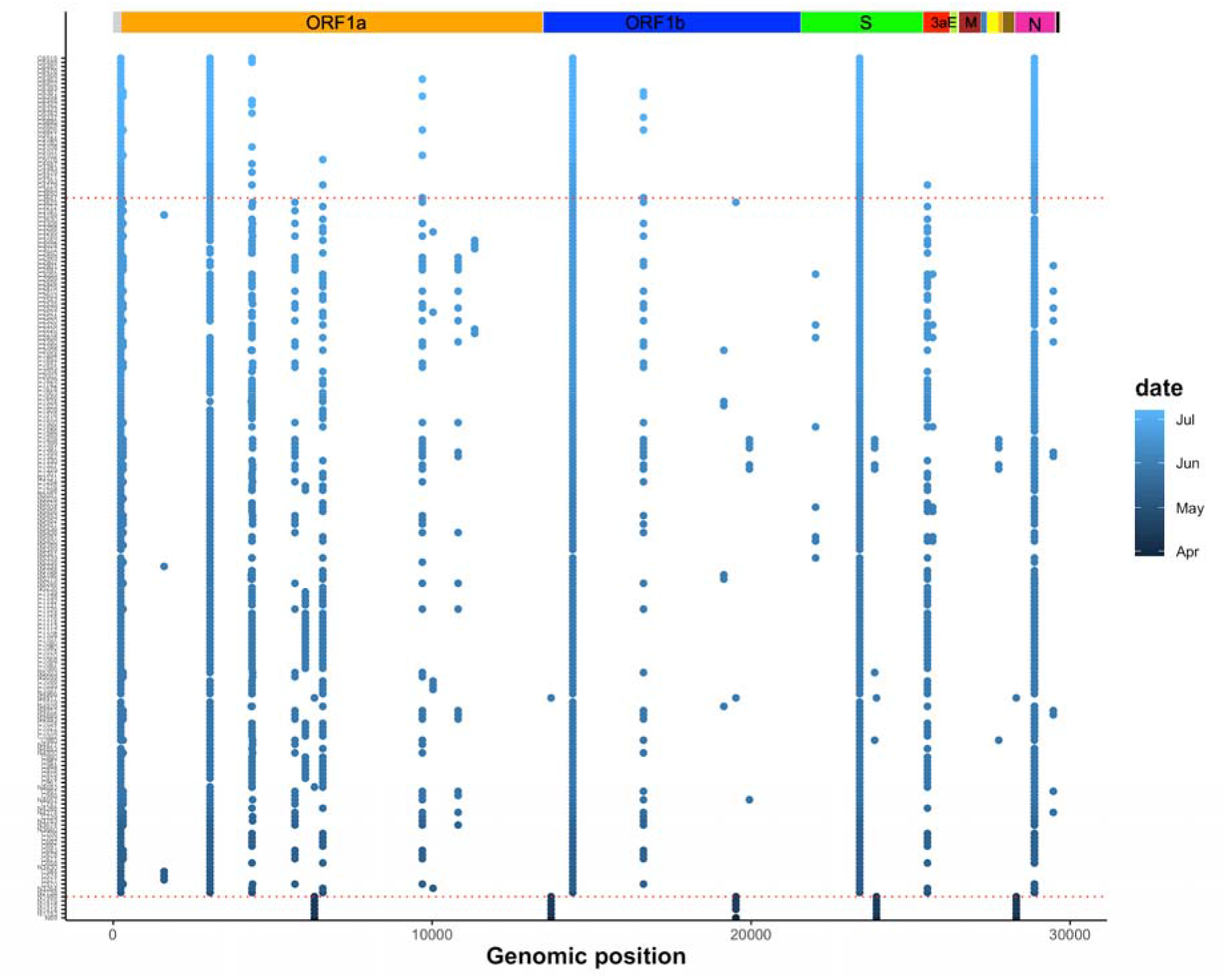
Mutational map across all the samples arranged with respect to date of collection. Horizontal red lines indicate the time at which major split were observed in the phylodynamic tree.

**Supplementary figure S4:**
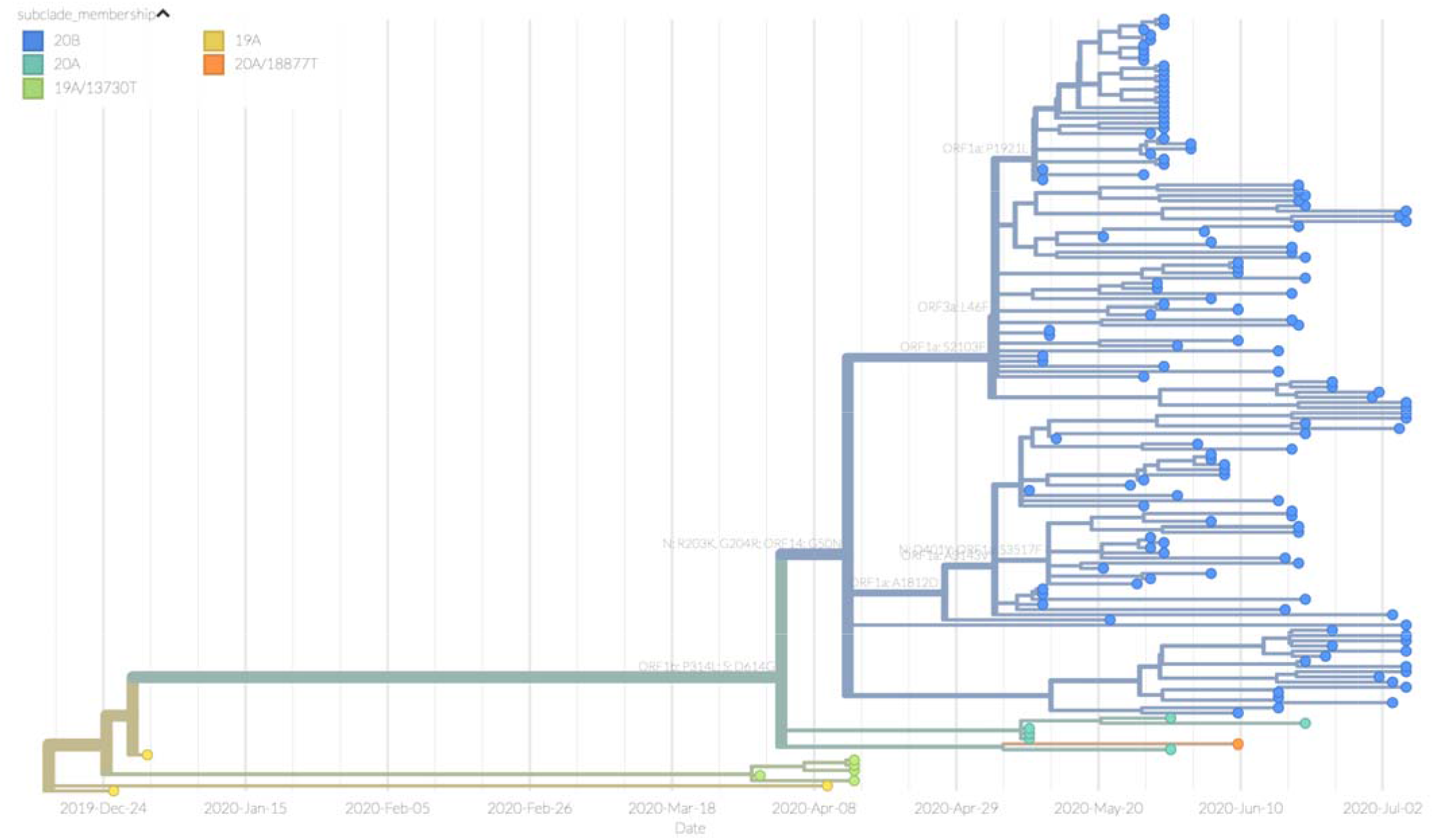
Time resolved phylogenetic tree created using Nextstrain.

**Supplementary figure S5:**
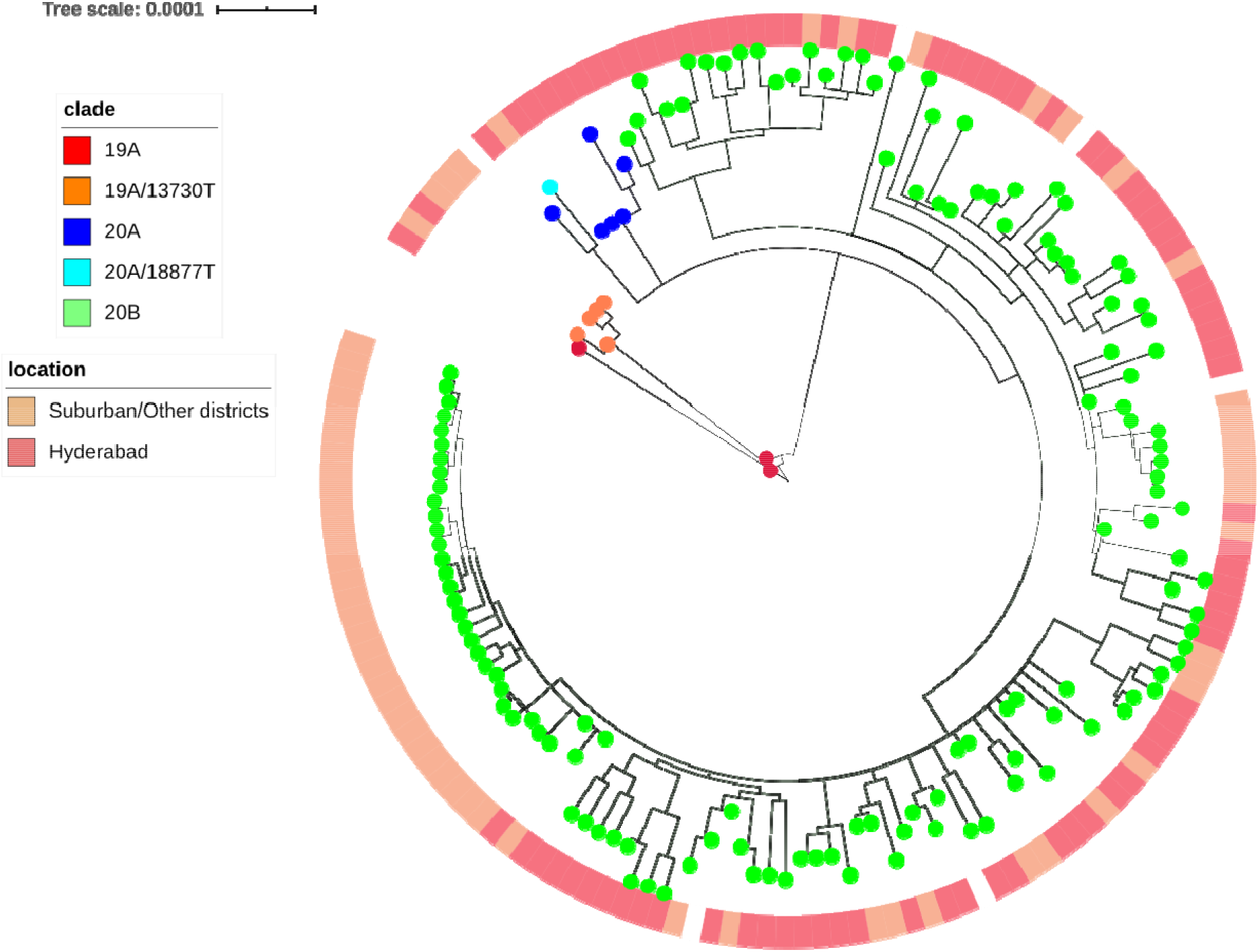
Phylogenetic tree with local distribution of samples.

**Supplementary figure S6:**
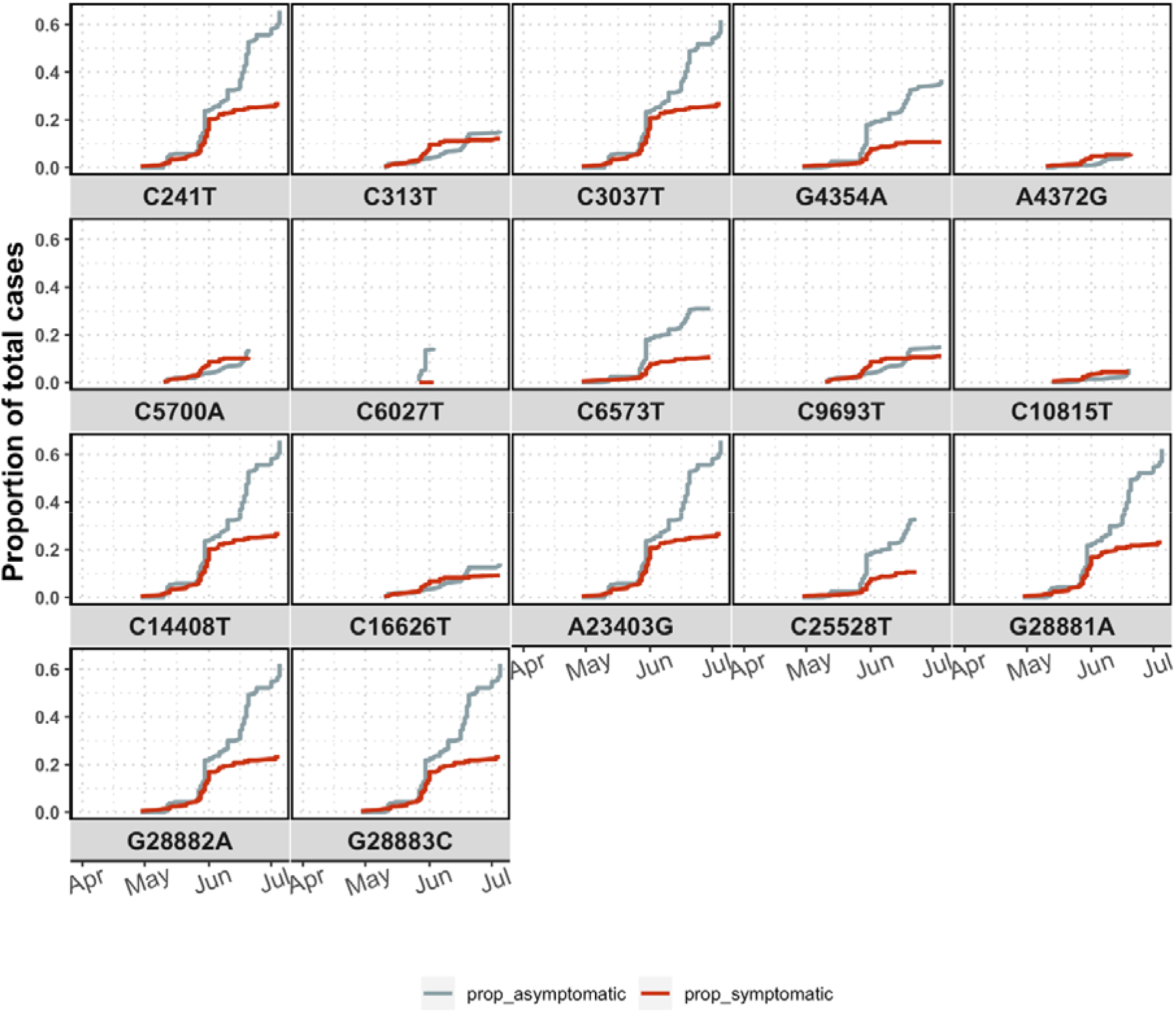
Cumulative frequency of samples showing asymptomatic and symptomatic behaviour for each of the 17 high frequency mutations.

**Supplementary data D1:**
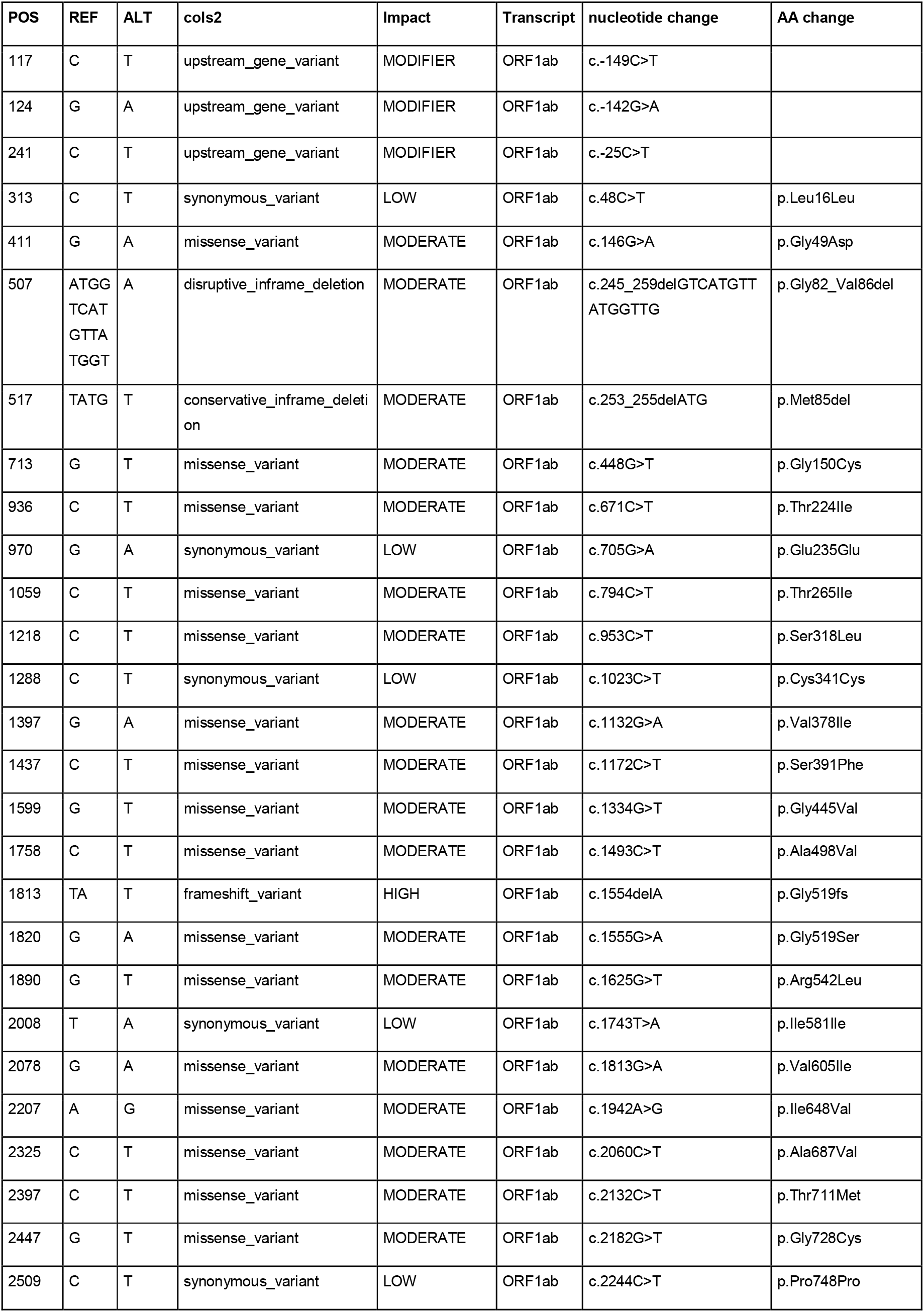
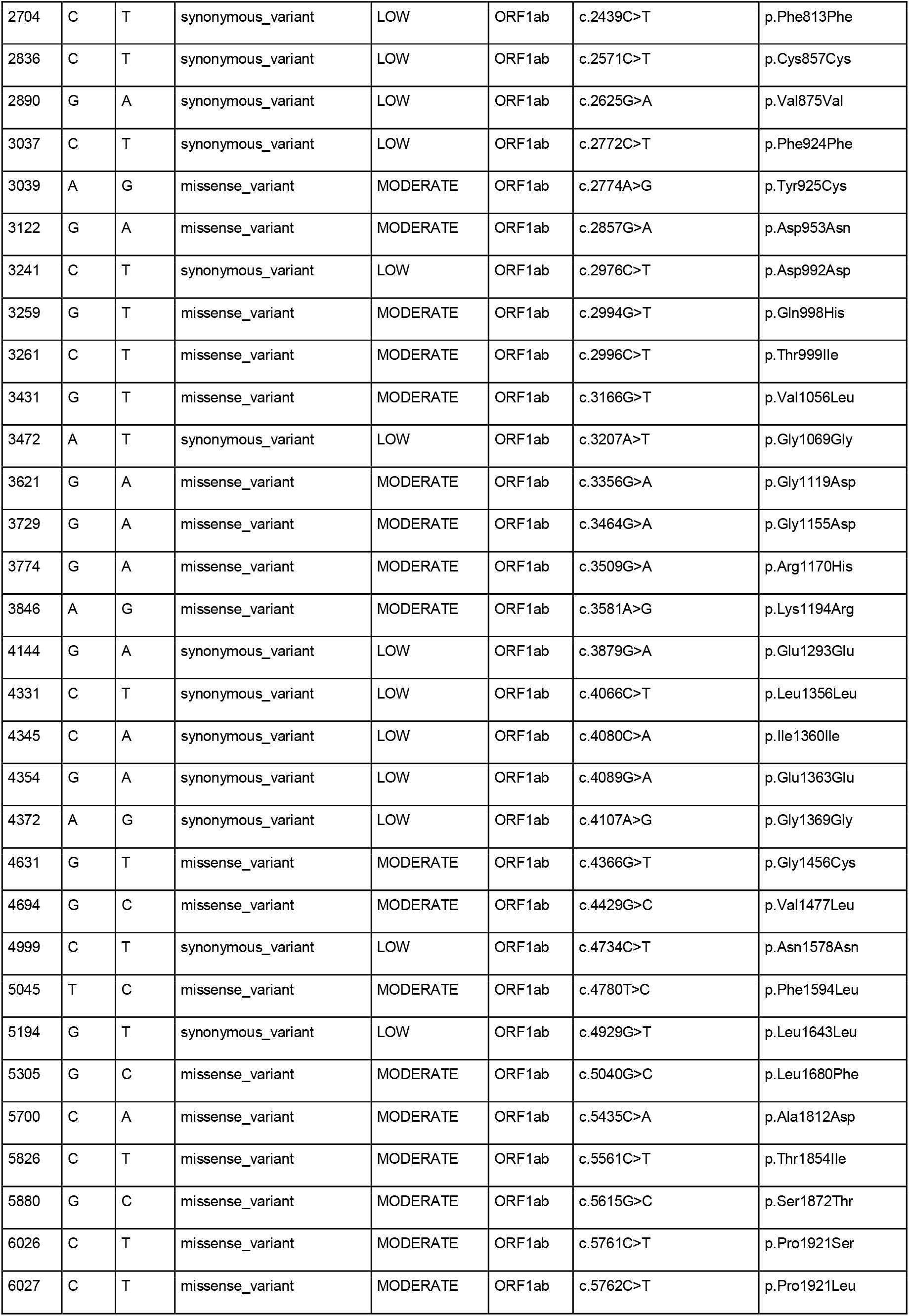
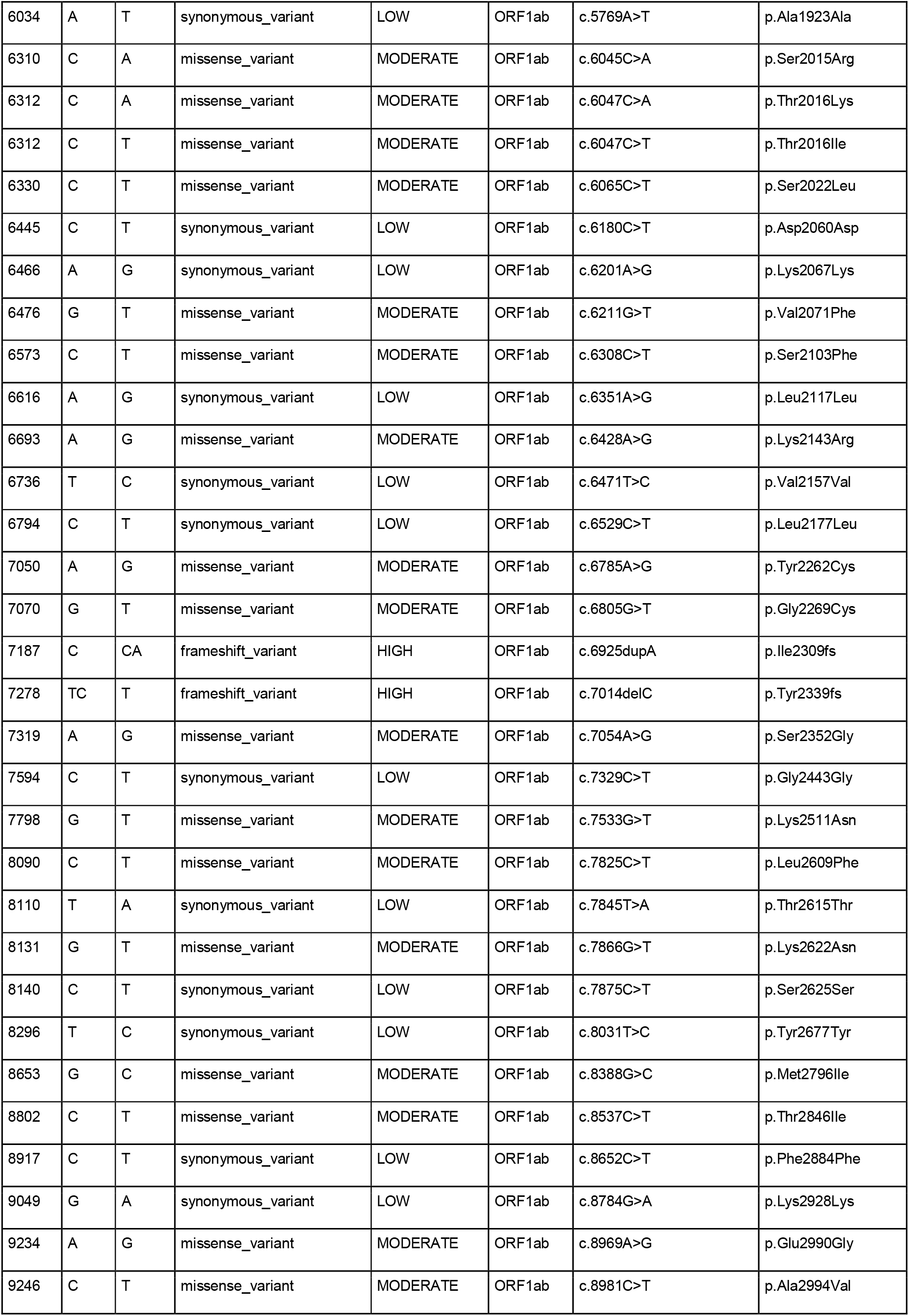
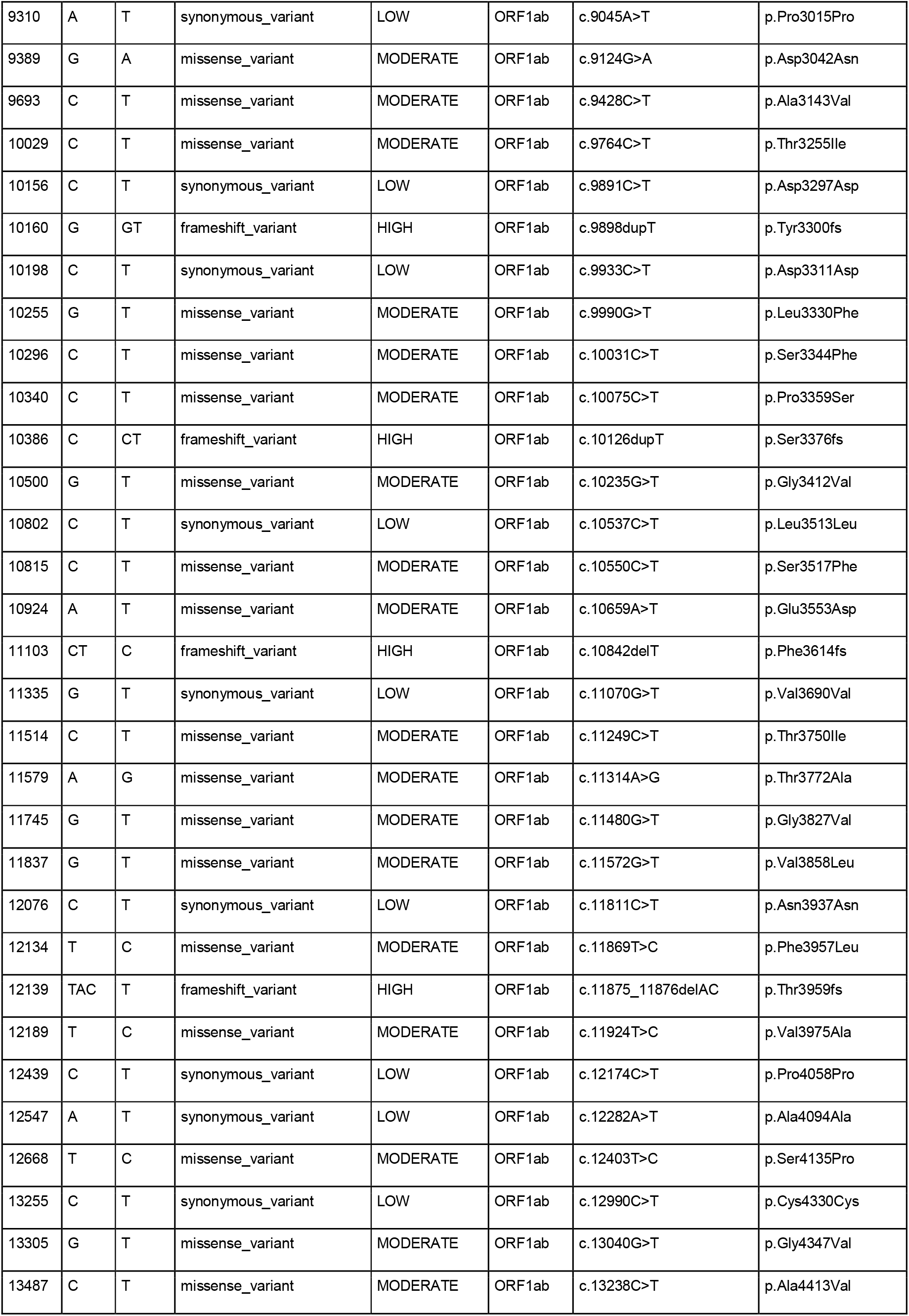
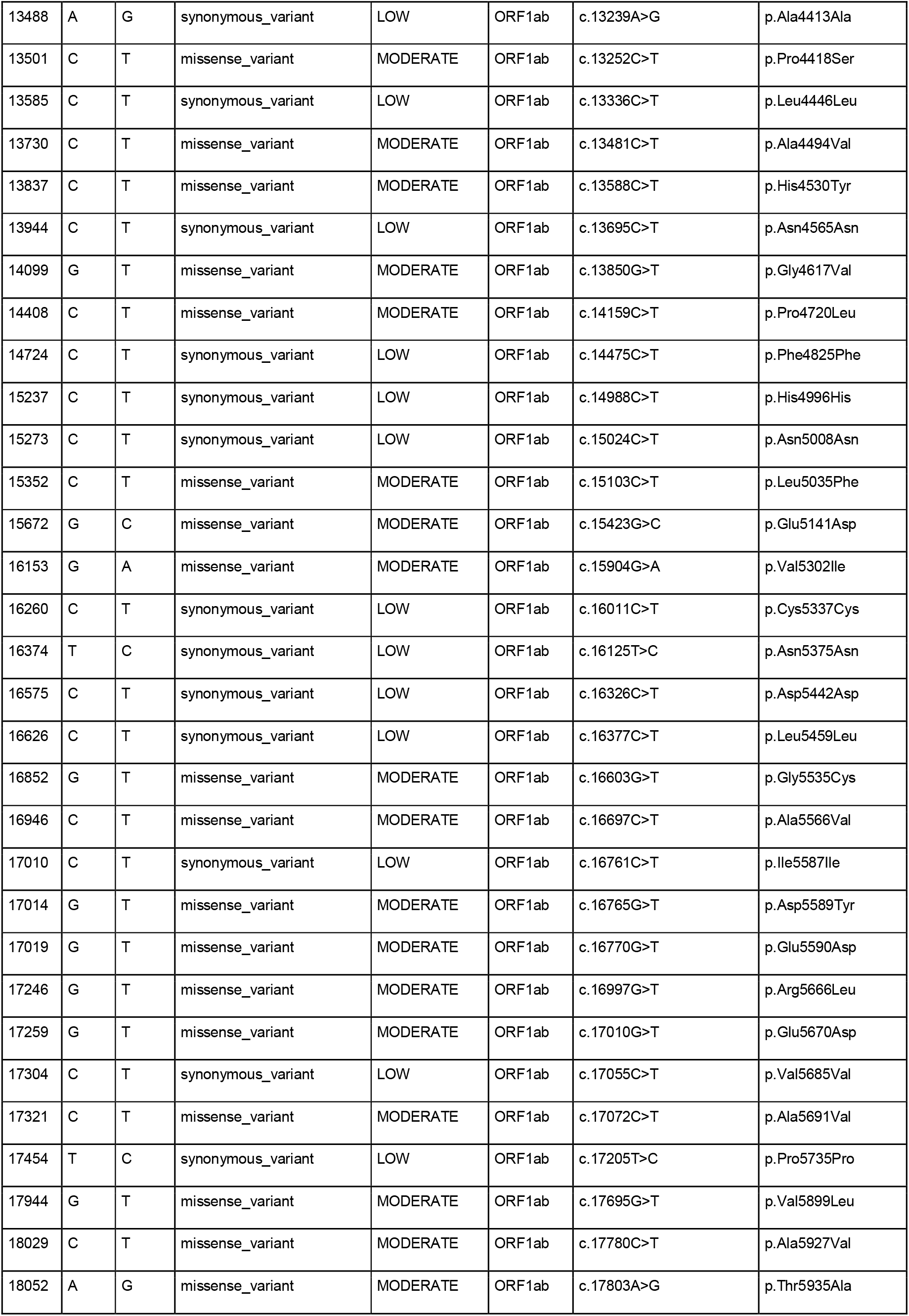
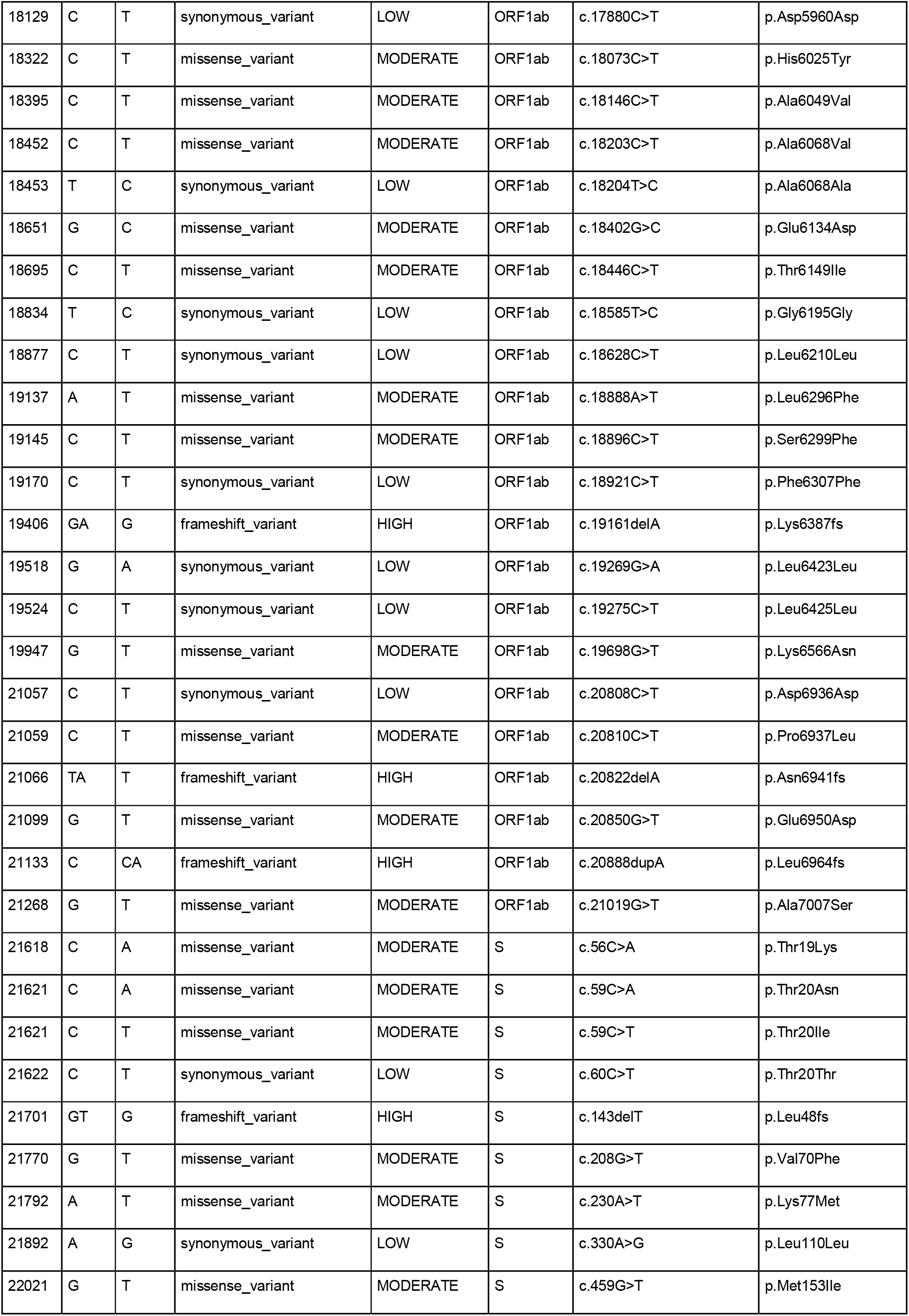
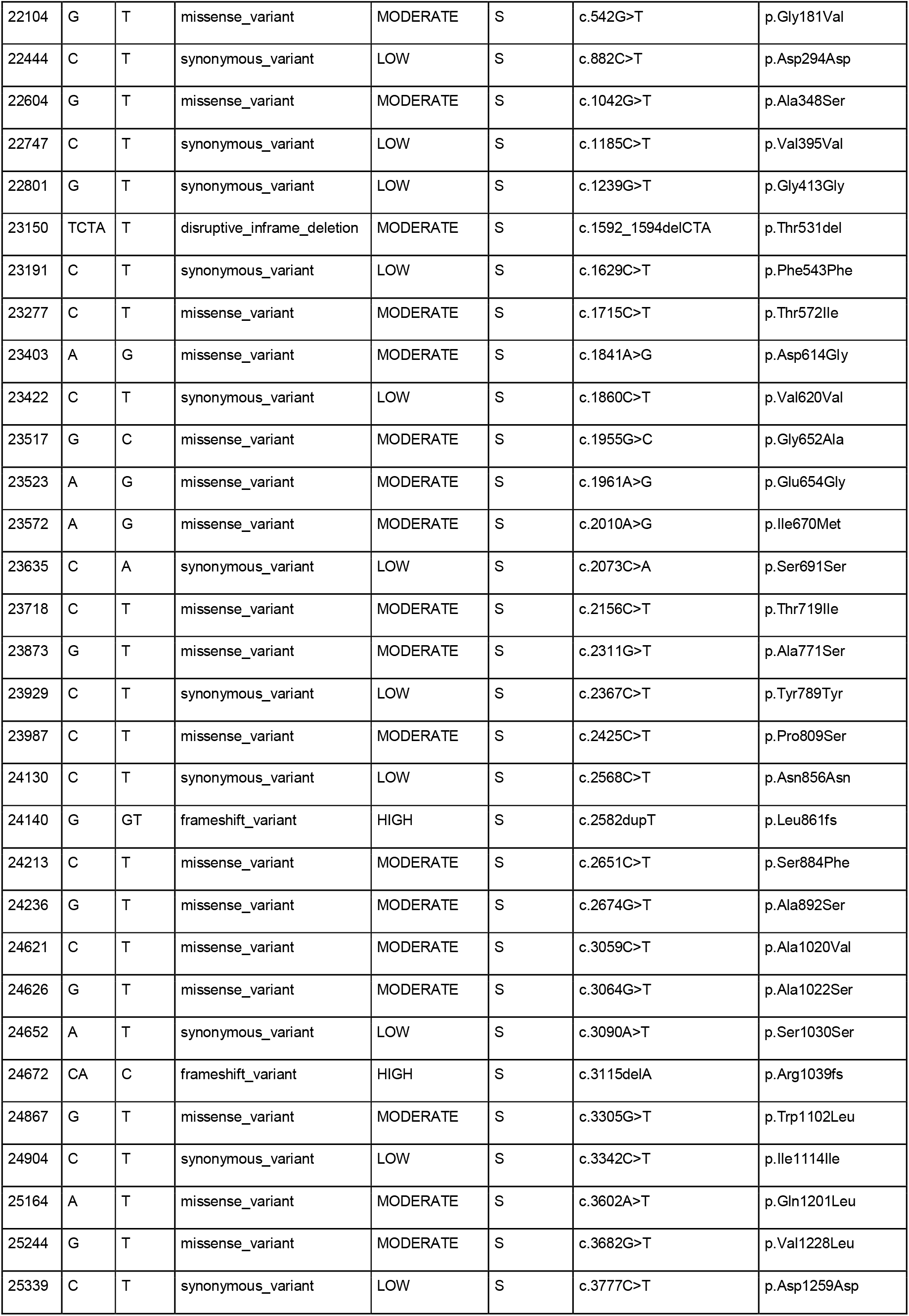
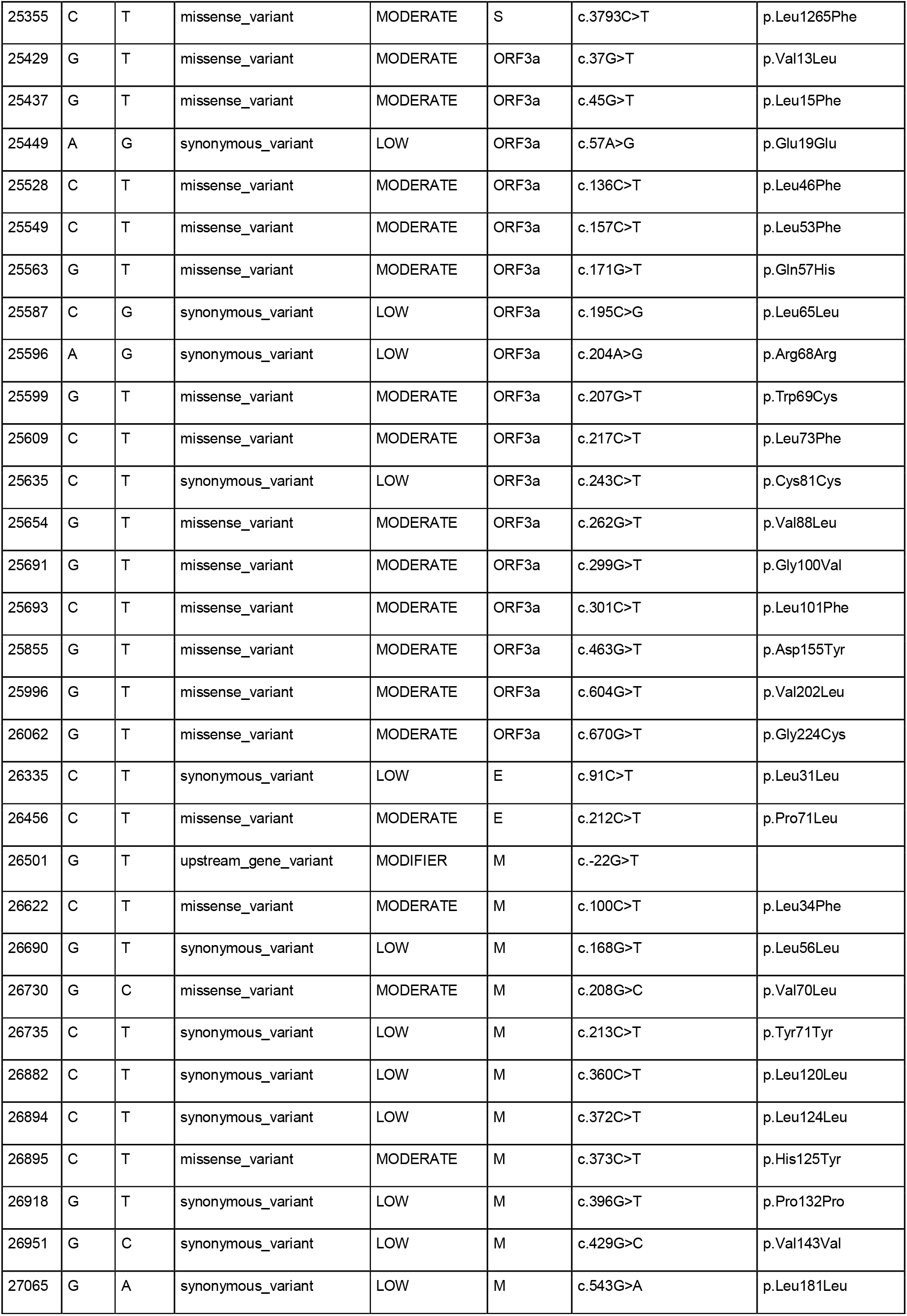
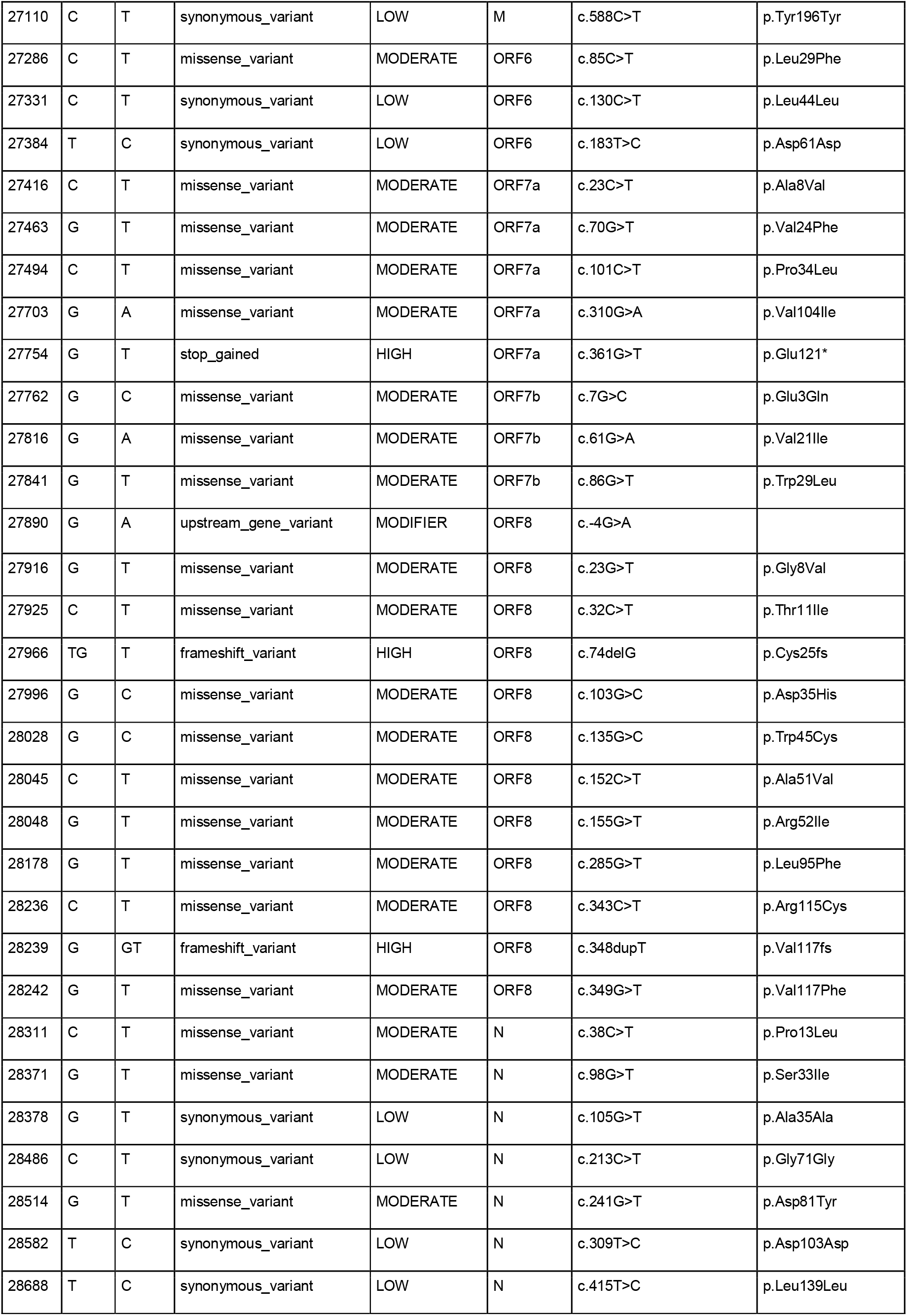
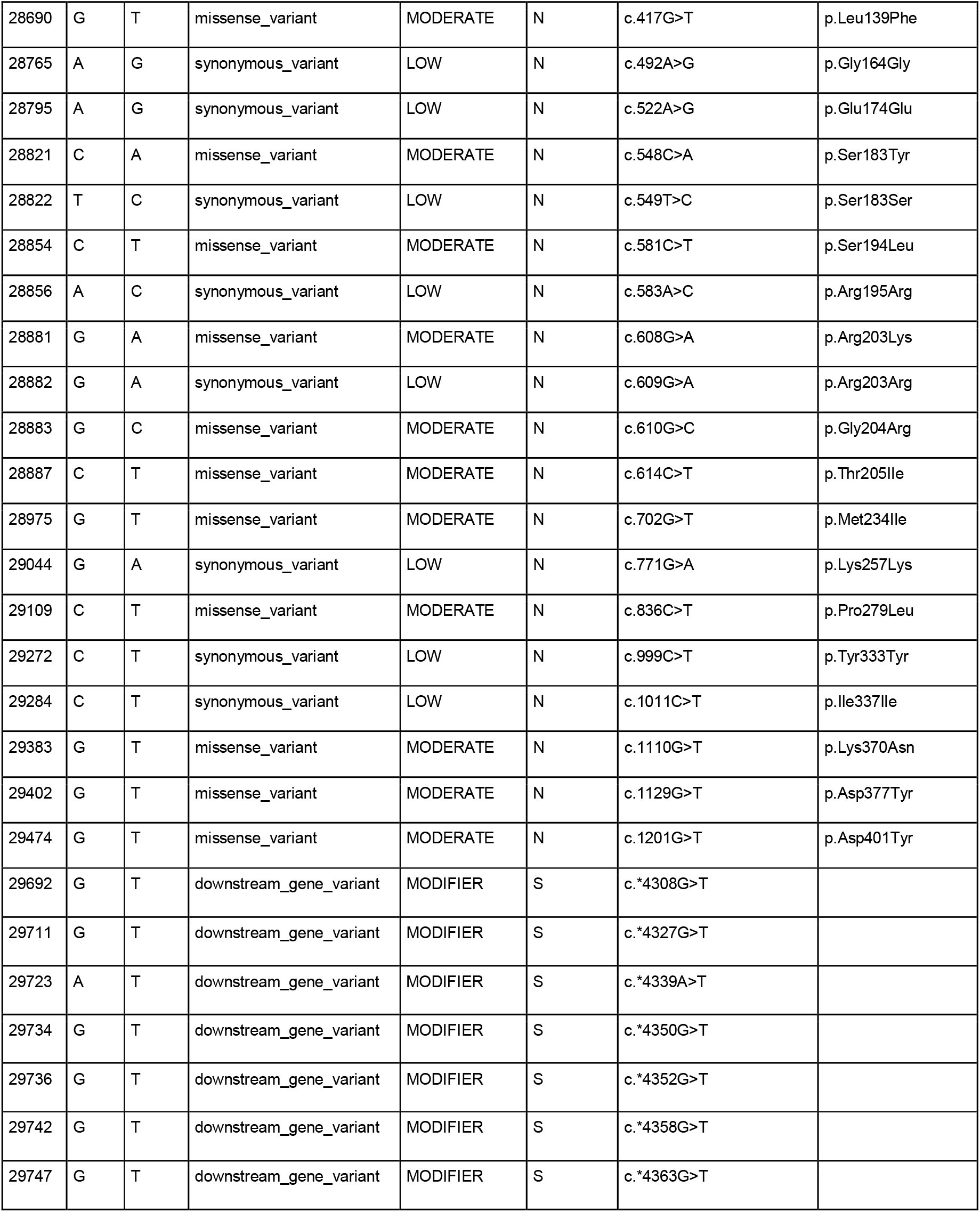
All the mutations identified in this study

